# Low Arousal Threshold Endotype and CPAP Adherence: Insights from a Large, Diverse Clinical Cohort

**DOI:** 10.1101/2025.10.17.25337940

**Authors:** Christopher N. Schmickl, Christian D. Harding, Athiwat Tripipitsiriwat, Jeremy E. Orr, Andrey Zinchuk, Pamela DeYoung, Stacie Moore, Robert L. Owens, Scott A. Sands, Atul Malhotra

## Abstract

**Background:** A low respiratory arousal threshold (ArTH) has been linked to reduced continuous positive airway pressure (CPAP) adherence in obstructive sleep apnea (OSA) via a multi-trait model developed in the RICCADSA trial. Our objective was to validate the prior model in a large, real-world cohort and explore alternative linear and non-linear approaches for predicting CPAP adherence.

**Research Question:** Does a previously derived multi-trait model linking low ArTH to poor CPAP adherence remain valid in a diverse real-world population and do alternative linear or non-linear approaches offer improved predictive performance?

**Methods:** Adults with OSA from the SNOOzzzE-cohort who initiated CPAP within 1 year of in-lab polysomnography (2017–2019) were included. Pathophysiological traits (Vpassive, Vactive, loop gain, ArTH, ventilatory response to arousal) were estimated from polysomnography. Poor (vs good) adherence was defined as ≤2.48h/night at month 1 (1^st^ sample quartile). The prior model’s performance was evaluated via the area under the curve (AUC) and adherence comparisons. Secondary analyses tested a *de novo* linear regression and a latent profile analysis (LPA)–derived trait-cluster model.

**Results:** Among 744 participants (45% women, 47% non-White), median CPAP adherence was 4.8 h/night at 1 month. The prior model’s AUC was 0.51 (95%-CI 0.46–0.55), with no usage differences between predicted poor vs good adherers. A new linear model overfit in training (AUC=0.85) but failed in testing (AUC=0.55). LPA identified a “Low ArTH Driven” cluster with persistently lower adherence at months 2–3 (P<.05) and a “Low ArTH & High loop gain” cluster whose usage stabilized after month 1.

**Conclusions:** The prior model did not generalize to this diverse clinical cohort. LPA identified a “Low ArTH Driven” endotype with persistently low CPAP adherence, suggesting potential for targeted interventions pending external validation.

## INTRODUCTION

Obstructive sleep apnea (OSA) affects ∼10% of US adults and is characterized by recurrent upper airway collapse during sleep, leading to transient hypoxemia and arousals.^1–3^ Untreated OSA is associated with many serious adverse health outcomes, including neurocognitive (e.g. excessive sleepiness^4,5^, motor-vehicle accidents^6,7^) and cardiovascular (e.g. hypertension^8,9^, coronary artery disease^10^ [CAD], mortality^11^) sequelae. While continuous positive airway pressure (CPAP) is the first-line treatment and highly efficacious in preventing upper airway collapse,^12^ adherence is variable, limiting real-world effectiveness.^13–16^

CPAP adherence is influenced by a range of biopsychosocial factors—including device comfort, disease awareness, and partner support.^17^ Despite advances such as improved mask designs, humidification, and patient engagement tools, adherence remains suboptimal.^14,18^ A promising avenue to improve adherence lies in leveraging our growing understanding of OSA as a heterogeneous disorder driven by interactions of multiple underlying mechanisms or “endotypes”.^19–21^

One such trait, a low respiratory arousal threshold (ArTH; a tendency to wake easily from a respiratory stimulus), contributes to OSA pathogenesis in ∼40% of patients,^19,22,23^ and, when measured from polysomnography signals and included in a multi-trait model, ArTH was associated with reduced long-term CPAP adherence in the RICCADSA trial.^24^ This model may help identify patients at risk for poor adherence and enable early, targeted interventions (e.g., hypnotics).^25^ While there is some debate about the value of estimating traits from polysomnography,^26,27^ one form of validation is to demonstrate clinical relevance. Thus, in line with recent calls to translate OSA endotyping into clinical care,^21^ our primary objective was to validate the OSA trait adherence model from the RICCADSA study (from now on RICCADSA-model) in a large, real-world clinical cohort. We hypothesized that model performance would be similar as before. Our secondary aim was to assess whether there was a relationship between ArTH and adherence using 1) a *de-novo* regression model with traits as continuous variables and 2) patients categorized into groups using the traits in latent profile analysis.

## METHODS

### Study Population

The San Diego Multi-Outcome OSA Endophenotype (SNOOzzzE) cohort includes 3,319 consecutive adults who underwent a clinical in-laboratory polysomnography at the UCSD sleep clinic between 1/2017-12/2019 (IRB #182136). For this analysis we included patients with OSA (i.e., apnea hypopnea index >5/h), whose in-laboratory polysomnography allowed for assessment of physiological traits (see below), and who newly initiated CPAP (using fixed pressure or auto-titrating mode) within 1 year of their polysomnography based on information in the cloud portals (i.e., Airview or CareOrchestrator) of the major CPAP manufacturers. We excluded patients who did not attempt CPAP (i.e., no usage at all after set up), used a PAP device other than CPAP, or used supplemental oxygen.

### Measurements of Pathophysiological Traits (Predictors)

Mechanisms underlying OSA were estimated from raw signals obtained during non-rapid eye movement sleep portions of the in-laboratory polysomnography using published techniques.^28–31^ To assess upper airway collapsibility we quantified Vpassive, which estimates the flow as a percentage of eupneic ventilation (%eupnea) at eupneic drive (lower Vpassive denotes worse collapsibility). To assess responsiveness of upper airway dilator muscles, we quantified Vactive (flow at elevated drive, specifically the level of the arousal threshold). Vcomp (Vactive-Vpassive) reflects the change in flow from passive to active conditions. Higher values of Vactive and Vcomp denote better upper airway dilator muscle function. Ventilatory instability (“loop gain”) was estimated as the magnitude of the ventilatory drive response to a prior reduction in ventilation in the time frame of 1 minute (loop gain 1, LG1) and the natural frequency (loop gain n, LGn; range 0 to infinity, higher levels of loop gain denote more instability). The arousal threshold (ArTH) was estimated as the level of respiratory drive causing arousals (lower values denoting easier arousability). Furthermore, the ventilatory response to arousal (VRA) was estimated as the ventilatory overshoot (%eupnea) in response to arousals (greater overshoots may worsen sleep apnea)^32,33^. Vpassive (Vpassive^T^ = 100-100*sqrt[1-Vpassive/100]) and Arousal Threshold (ArTH^T^ = 100+100*sqrt[ArTH/100-1]) were transformed for normality as in prior studies with transformed values used throughout.^34,35^

### CPAP adherence (Outcome)

Adherence was defined as average usage per night, and abstracted manually from device manufacturer’s cloud portals for months 1, 2 and 3 following set up with CPAP. As before, the lowest quartile of CPAP usage was classified as “poor” adherence (vs all others as “good”). Patients who had used CPAP during month 1 but lacked adherence data for months 2 and/or 3, were assumed to have discontinued CPAP and imputed as zero use for those months.

### Validation of the RICCADSA-Model

As in the development cohort, traits were mean-centered (using means from RICCADSA: ArTH^T^ 143.5 %eupnea, LG1 0.67, Vpassive^T^ 143.5 %eupnea, Vcomp 0.43 %eupnea) and outliers were identified and handled as before (three Vcomp values were trimmed to 80.7%). Then, for each participant predicted adherence (ŷ) was calculated using the following formula: ŷ = 3.722 – 0.012 x TimeMonths + 0.026 x Arth^T^ – 0.496 x LG1 + 0.019 * Vpassive^T^ – 0.007 x Vcomp – 0.000475 x Vcomp^2^. Replicating the prior analytical approach, model performance was assessed a) via the area under the receiver operating characteristic curve (AUC) for identifying “poor” adherers at 1 month (see above), and b) by comparing actual median CPAP adherence in patients *predicted* as “poor” vs “good” adherers for months 1-3 using Wilcoxon rank sum tests (using the lowest quartile of ŷ to denote patients who were *predicted* to be “poor” adherers).

To assess robustness of results, sensitivity analyses repeated assessments in various subsets that resembled the RICCADSA cohort more closely and/or had previously been found to show better model performance (patients with AHI>15/h, patients with coronary-artery disease [CAD], and men without REM-dominant OSA).

### Exploratory Analyses

Given suboptimal performance of the RICCADSA-model in the SNOOzzzE cohort, we explored two alternative approaches to assess whether the relationship between ArTH and CPAP adherence—rather than a specific prediction model—can be generalized across cohorts:

1) <u>Alternative linear model:</u> Using the polysomnography date of 1/19/2019 as the cut-off, we split the data into a training vs test set (60:40) and employed a similar approach as *Zinchuk et al.* to develop a *de novo* mixed effects linear regression model (with random intercepts and slopes) via backward selection based on the Akaike Information criterion (AIC) in the training set. ArTH^T^, LG1, Vpassive^T^, and Vcomp were forced into the model as main effects while the procedure considered 10 additional terms (four quadratic and six interaction terms) as candidates. Performance was assessed akin to the above.
2) <u>Alternative non-linear model:</u> Given the potentially complex interplay between traits that may be insufficiently captured by interaction terms in linear models, we assessed whether “trait clusters” (endotypes) identified via an unsupervised latent profile analysis (LPA) may help better identify patients at risk of low adherence. The LPA model was previously developed in 2,367 participants of the SNOOzzzE cohort following a standard approach^36,37^ and reported in abstract form;^38^ full methodological details are provided in **E-Appendix 1** of the online supplement. The model identified six distinct clusters (technically “profiles”), two of which contained a relatively low ArTH as a key pathophysiological feature. Cluster 2 is purely “Low ArTH-Driven”, while Cluster 5 is characterized by both a “Low ArTH & High LG”. The model can readily be applied to new trait data using a publicly available end-to-end pipeline (https://github.com/CSchmickl/SNOOzzzE_Endocluster_LPA_Pipeline), which was used to assign the participants in the current investigation to the six clusters based on posterior probabilities. To test our hypothesis that the two clusters including a low ArTH were associated with low CPAP usage, we compared median CPAP adherence in each of those clusters to the combined group of the remaining four clusters at each time point using Dunn’s test.

All analyses were performed in R (version 4.4.1; major packages: pROC, lmerTest), using P<.05 to denote statistical significance.

## RESULTS

The analysis included 744 patients who met all eligibility criteria (**Figure 1**). At one month median [interquartile, IQR] adherence was 4.8 [2.5, 6.4] h/night, which decreased slightly to 4.5 [1.2, 6.4] h/night at 3 months. The overall cohort was middle aged, and highly diverse including 45% women, 47% reporting non-White race, and 26% reporting Hispanic/Latino ethnicity. The 189 (25%) “poor” adherers with CPAP usage of ≤2.48 h/night at 1 month had generally similar characteristics as “good” adherers, but were more likely to report Black race and Hispanic/Latino ethnicity, and had slightly lower loop gain (**Table 1**).

**Figure 1.**
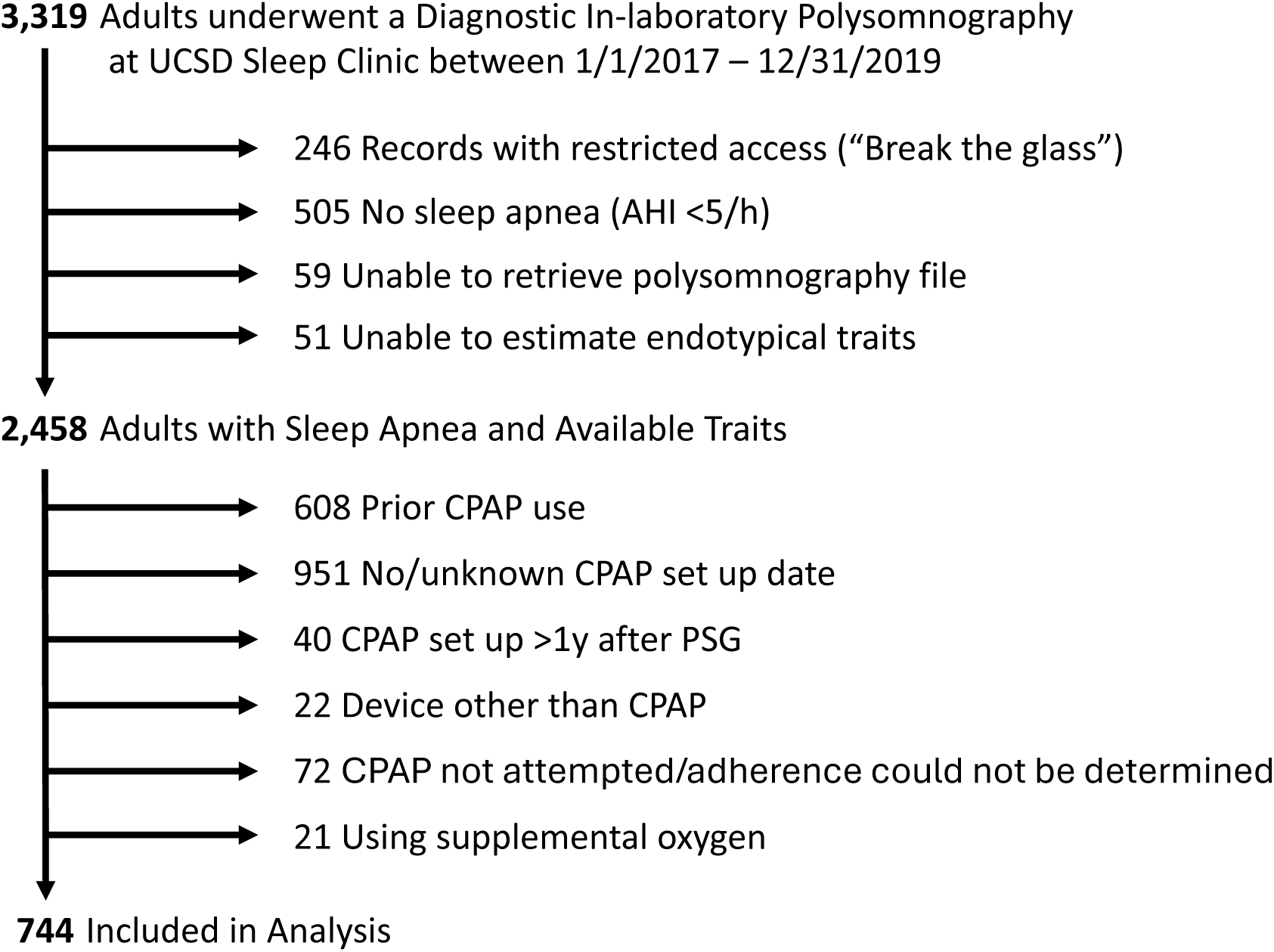
Study Flowchart.

**Table 1.** General Characteristics of Poor vs Good Adherers. Classified based on adherence at 1 month (“poor” adherers = CPAP use/night at or below the lowest quartile of adherence [≤2.48h/night]).

| Characteristic | Overall<br>N = 744<sup>1</sup> | Poor Adherers<br>N = 189<sup>1</sup> | Good Adherers<br>N = 555<sup>1</sup> | p-value<sup>2</sup> |
| --- | --- | --- | --- | --- |
| **Demographics** |  |  |  |  |
| Age, years | 56 (43, 66) | 55 (40, 64) | 57 (44, 67) | 0.051 |
| Female Sex | 333 (45%) | 85 (45%) | 248 (45%) | >0.9 |
| Race |  |  |  | 0.019 |
| Asian | 60 (8.1%) | 14 (7.4%) | 46 (8.3%) |  |
| Black | 49 (6.6%) | 17 (9.0%) | 32 (5.8%) |  |
| White | 396 (53%) | 84 (44%) | 312 (56%) |  |
| Other or Unknown | 239 (32%) | 74 (39%) | 165 (30%) |  |
| Hispanic or Latino Ethnicity | 190 (26%) | 61 (32%) | 129 (23%) | 0.014 |
| Body Mass Index (BMI), kg/m<sup>2</sup> | 32 (27, 38) | 33 (27, 40) | 32 (27, 38) | 0.3 |
| **Comorbidities** |  |  |  |  |
| Obesity (BMI >30 kg/m<sup>2</sup>) | 456 (61%) | 120 (63%) | 336 (61%) | 0.5 |
| Hypertension | 340 (46%) | 86 (46%) | 254 (46%) | >0.9 |
| CAD | 76 (10%) | 20 (11%) | 56 (10%) | 0.8 |
| **Sleep Characteristics & Symptoms** |  |  |  |  |
| Epworth Sleepiness Score (ESS) | 8.0 (4.0, 13.0) | 7.5 (4.0, 12.0) | 8.0 (5.0, 13.0) | 0.2 |
| Unknown | 114 | 35 | 79 |  |
| Excessive Sleepiness (ESS >10) | 240 (38%) | 52 (34%) | 188 (39%) | 0.2 |
| Unknown | 114 | 35 | 79 |  |
| N1, % | 10 (5, 20) | 10 (5, 17) | 10 (5, 20) | 0.081 |
| N2, % | 61 (51, 71) | 60 (51, 71) | 61 (51, 71) | 0.7 |
| N3, % | 9 (0, 20) | 10 (0, 22) | 8 (0, 18) | 0.2 |
| REM, % | 13 (5, 18) | 12 (4, 18) | 13 (5, 18) | >0.9 |
| AHI3A, events/h | 33 (20, 55) | 34 (20, 52) | 33 (20, 56) | 0.5 |
| OSA severity |  |  |  | 0.9 |
| Mild (AHI 5-15/h) | 105 (14%) | 27 (14%) | 78 (14%) |  |
| Moderate (AHI 15-30/h) | 226 (30%) | 56 (30%) | 170 (31%) |  |
| Severe (AHI $\geq 30$/h) | 413 (56%) | 106 (56%) | 307 (55%) | |
| % Hypopneas | 74 (49, 91) | 78 (56, 91) | 74 (48, 91) | 0.2 |
| Central AHI, events/h | 0.01 (0.00, 0.06) | 0.02 (0.00, 0.06) | 0.01 (0.00, 0.07) | 0.5 |
| NREM-AHI, events/h | 32 (16, 54) | 32 (16, 51) | 32 (17, 55) | 0.4 |
| REM-AHI, events/h | 45 (24, 62) | 47 (19, 62) | 45 (24, 62) | >0.9 |
| Unknown | 152 | 40 | 112 |  |
| Hypoxic Burden, %min/h | 62 (36, 112) | 65 (35, 115) | 62 (36, 112) | 0.8 |
| T90, % | 3 (1, 9) | 3 (1, 9) | 3 (1, 10) | >0.9 |
| SpO<sub>2</sub> nadir, % | 81 (75, 85) | 80 (74, 85) | 81 (76, 85) | 0.3 |
| **OSA Pathophysiological Traits** |  |  |  |  |
| Vpassive<sup>T</sup>, %eupnea | 68 (55, 77) | 70 (56, 77) | 68 (55, 77) | 0.2 |
| ArTH<sup>T</sup>, %eupnea | 144 (127, 161) | 142 (124, 159) | 144 (127, 161) | 0.5 |
| Loop Gain n | 0.41 (0.34, 0.48) | 0.40 (0.33, 0.48) | 0.42 (0.35, 0.49) | 0.029 |
| Loop Gain 1 | 0.60 (0.49, 0.71) | 0.57 (0.47, 0.68) | 0.60 (0.50, 0.73) | 0.011 |
| Vcomp, %eupnea | 4 (0, 10) | 4 (1, 11) | 4 (0, 10) | 0.3 |
| VRA, %eupnea | 33 (22, 54) | 34 (22, 60) | 33 (22, 52) | 0.7 |
| **CPAP Usage** |  |  |  |  |
| PAP use/night, Month 1, h | 4.80 (2.48, 6.37) | 1.12 (0.38, 1.87) | 5.48 (4.40, 6.78) | - |
| PAP use/night, Month 2, h | 4.73 (1.98, 6.36) | 0.27 (0.00, 1.60) | 5.45 (4.07, 6.90) | - |
| PAP use/night, Month 3, h | 4.48 (1.17, 6.36) | 0.08 (0.00, 1.65) | 5.35 (3.78, 6.72) | - |

<sup>1</sup>Median (Q1, Q3); n (%)
<sup>2</sup>Wilcoxon rank sum test; Chi-squared test

When the RICCADSA-model was applied to the SNOOzzzE cohort, its AUC was 0.51 (bootstrapped 95%-confidence interval, CI 0.46 to 0.55), compared to an AUC of 0.65 reported for the development cohort (RICCADSA).^24^ Median CPAP adherence during months 1-3 was similar in those *predicted* to be poor vs good adherers (P>0.3; **Figure 2**). Model performance was similarly poor in the subgroups of patients with AHI>15/h (n=639), CAD (n=76), or men without REM-dominant OSA (n=352; data not shown).

**Figure 2.**
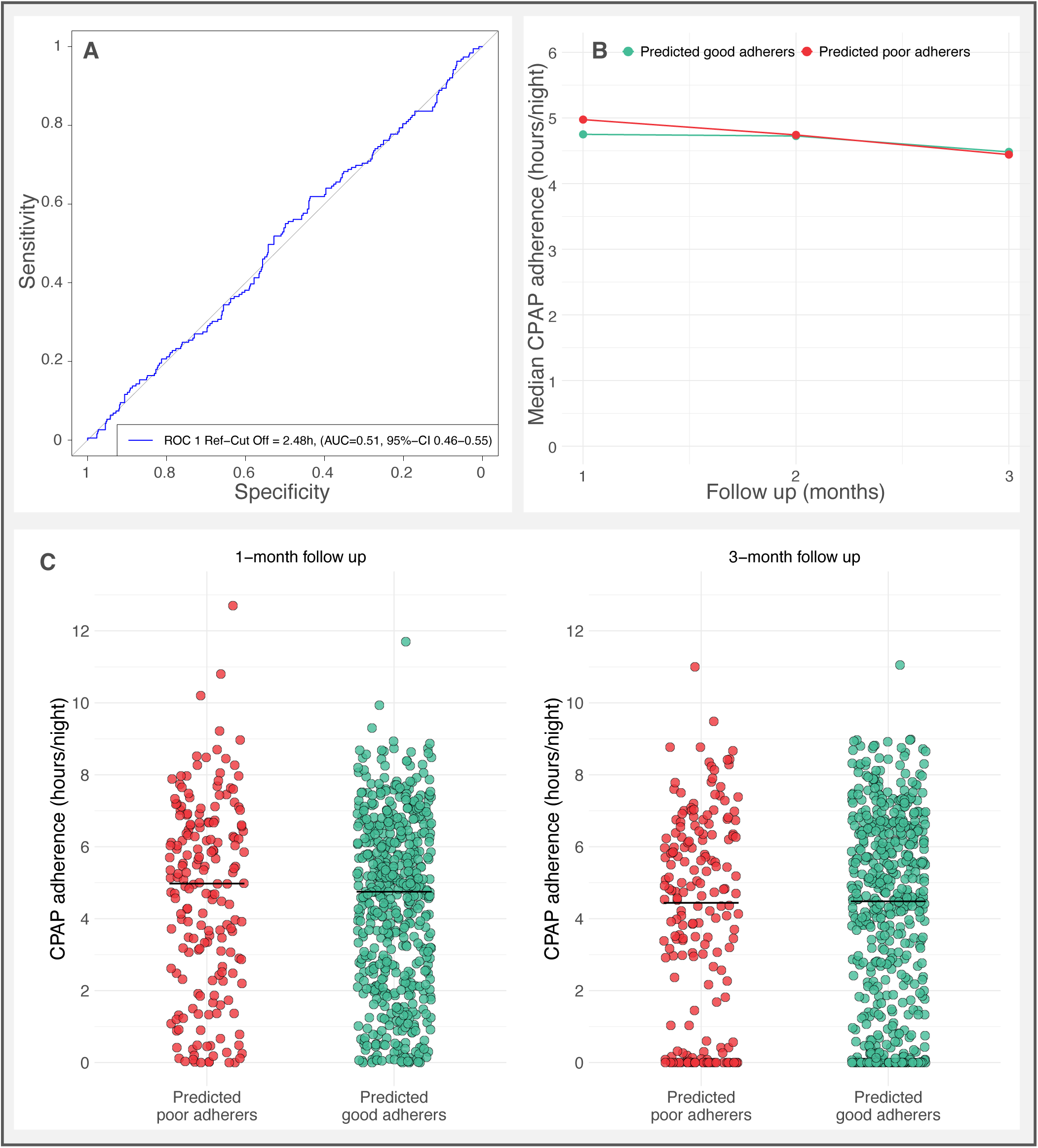
Performance of the RICCADSA-Model in the SNOOzzzE cohort. Panel A shows the AUC for adherence at 1 month. Panel B shows the median actual CPAP adherence for patients predicted to be poor (red) vs good (green) adherers across months 1-3. Differences between groups were non-significant at each time point (P>0.3). Panel C shows the distributions and medians (bars) of CPAP adherence among those with predicted poor vs good adherence at the 1– and 3-months follow-up.

The optimal linear trait model newly developed in the training set of the SNOOzzzE cohort included one interaction between loop gain and Vpassive (higher loop gain being associated with greater adherence, especially in patients with mild collapsibility; **E-Table 1**). Notably, ArTH was not a significant predictor of CPAP adherence over the 3-month follow up. This *de novo* model performed well in the training set, but not in the test set (AUCTrain 0.85 [95%-CI 0.82-0.88] vs AUCTest 0.55 [95%-CI 0.48-0.63]). Results were similar in sensitivity analyses when including demographics (i.e., age, sex, race, and ethnicity) to the model (**E-Figure 1**).

When applying the LPA model to the 744 patients in this study, 24% (180) were assigned to the “Low ArTH Driven” cluster and 11% (81) to the “Low ArTH & High LG” cluster (**Figure 3**). Compared to patients in the other four clusters, those in the two low ArTH clusters were younger, more often female, and had milder OSA (**E-Table 2**). In the non-ArTH clusters, median [IQR] CPAP adherence declined from 5.0 [2.4, 6.6] h/night at 1 month to 4.9 [2.1, 6.5] h/night at 2 months and 4.6 [1.3, 6.6] h/night at 3 months. In comparison, median adherence in the “Low ArTH Driven” cluster was 0.5 h/night lower at 1 month (P=.19) and 0.7 h/night lower at months 2 and 3 (P<.05; **Figure 3**). Of note, median adherence in the “Low ArTH & High LG” cluster started out similarly low as in the “Low ArTH Driven” cluster at 1 month (4.6 [2.5, 5.8] vs 4.5 [2.5, 6.1] h/night), but in months 2 and 3 appeared more similar to the non-ArTH clusters.

**Figure 3.**
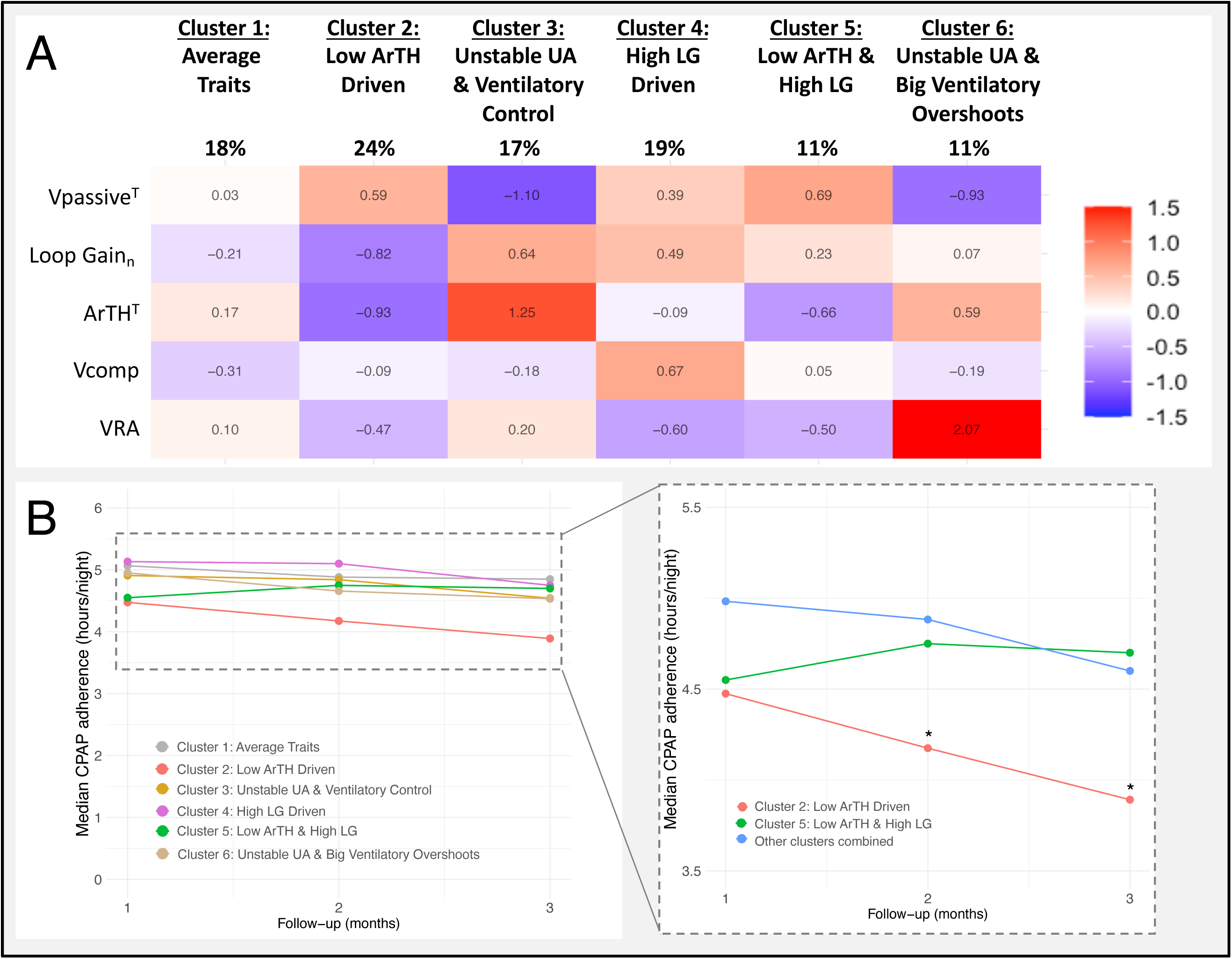
Pathophysiological Trait Clusters (Endotypes). Panel A: Prevalence and trait profiles of the six clusters, shown as a heatmap of standardized values (z-scores; red = positive, blue = negative). Conceptually, for loop gain (LG) and ventilatory response to arousal (VRA) higher z-scores indicate contribution to OSA pathogenesis, whereas for Vpassive, arousal threshold (ArTH), and Vcomp lower z-scores indicate contribution. Panel B: Median CPAP adherence across months 1-3 by cluster. Adherence was consistently lowest in the “Low ArTH Driven” cluster (asterisks indicate P<.05 vs non-ArTH clusters for months 2 and 3). The “Low ArTH & High LG” cluster started out at 1 month with similarly low adherence, but then aligned with the non-ArTH clusters during months 2 and 3. For a detailed boxplot see **E-Figure 2**. Abbreviations: UA upper airway.

## DISCUSSION

In this large, diverse clinical cohort of adults with OSA initiating CPAP therapy, early adherence was comparable to national benchmarks^15,16^ and as expected declined slightly over the first three months,^24^ with substantial variability between individuals. Notably, self-reported Black race and Hispanic/Latino ethnicity were associated with lower CPAP adherence, consistent with prior reports of disparities in CPAP use.^39,40^ But the main finding was that the RICCADSA model^24^ failed to validate in our cohort, performing no better than chance. Importantly, the LPA model suggests that low ArTH is an important determinant of CPAP adherence, but its effect is modified by other traits in ways that may not be sufficiently captured by standard regression models.

Possible reasons for the poor predictive performance of the RICCADSA-model in the current study include differences in signal quality and scoring that could potentially affect estimates of traits (e.g., Chicago criteria for RICCADSA vs AHI 3% or arousal for SNOOzzzE),^41^ patient populations and settings (e.g., mostly white males from Sweden in a trial setting in RICCADSA vs a diverse, insurance-bound, real world US-cohort in SNOOzzzE). The latter could be tested by applying the RICCADSA-model to other trial populations with similar characteristics and if performing well could still be useful to optimize adherence for future research studies. Another possible explanation for the poor model performance in in this investigation is model overfitting (N was 249 in RICCADSA; no separate validation cohort, with cross-validation potentially providing some false reassurance). To assess this hypothesis, we developed a new linear multi-trait model, which performed well in the training set but poorly in the test set—again suggesting overfitting and underscoring the importance of independent validation.

We then considered whether the effect of the ArTH on adherence might be modified by other traits in complex, non-linear ways. Indeed, an unsupervised latent profile analysis (LPA) identified two clusters containing a low ArTH but with opposite adherence trajectories: both clusters started out with relatively low CPAP adherence at 1 month; but in patients in whom the low ArTH was the primary driver, adherence continued to decline, whereas in those who had a low ArTH and high loop gain CPAP usage stabilized or improved after month 1. Mechanistically, high loop gain reflects a greater increase in respiratory drive for a given drop in ventilation, potentially making arousals more likely and initially contributing to intolerance. However, both loop gain and ArTH tend to decrease with CPAP use,^42–44^ which may explain the improvement in the “Low ArTH & High LG” cluster versus the continued decline in the “Low ArTH Driven” cluster. The reduction in CPAP usage in the “Low ArTH Driven” cluster represents a clinically meaningful difference,^45^ and its association with self-reported Black race may point to an actionable target for addressing OSA disparities. Our results are also consistent with a recent big-data analysis of CPAP usage patterns during the first 90 days in nearly 800,000 patients with OSA, which found that young women—who were disproportionately represented in the “Low ArTH Driven” cluster—exhibited the steepest decline in CPAP use compared to other demographic groups.^46^

Considering that traits do not exist in isolation (i.e., in a given individual all traits always co-exist and exert pathophysiological effects jointly), these findings also illustrate how LPA-based clustering may serve as a bridge from individual pathophysiological traits to actual endotypes—subgroups defined by a shared underlying mechanism^47^—which may ultimately guide personalized treatment strategies. However, external validation and impact analyses will be essential before clinical application.^21^

While cluster analysis has been widely used to identify symptom subtypes and provided many important insights,^21,48–50^ to our knowledge there has been only one prior study by *Cheng et al.* applying this approach to pathophysiological traits to assess a potential link with OSA symptoms.^51^ In a prospective Taiwanese cohort, latent *class* analysis based on four dichotomized traits (UA collapsibility, ArTH, LG, UA muscle compensation) identified 3 clusters: a high collapsibility/loop gain cluster (similar to our cluster 3), a single low ArTH cluster, and a low compensation cluster which Cheng et al. hypothesized to be idiosyncratic to the Taiwanese population. Trait dichotomization likely produced overly crude subgroups, and an additional latent *profile* analysis (LPA) allowing for inclusion of traits into the model as—more informative—continuous variables reportedly suggested a four cluster solution, but two clusters were only gradually different (low vs very low compensation, i.e., “salsa effect”) rather than being biologically distinct.^36^ The relationship between clusters and CPAP adherence was not assessed. Differences in trait clusters identified in both studies likely relate to differences in methodology (e.g., use of AHI4 to estimate traits^52^ and not including VRA in the prior study), population (diverse US-based vs Taiwanese), and sample size (N >1,500 vs ∼500).

The strengths of our study include its large, diverse clinical sample, assessment of a previously published model of physiological traits and adherence, and the rigorous cluster analysis approach to identify latent physiological profiles of OSA. Moreover, while the prior symptom cluster literature is characterized by each study developing a new model—important to assess generalizability of identified clusters but precluding validation of the relationship between the clusters of a *specific* model and outcomes, and thus translation into practice—we made our trait-cluster model publicly available so it can readily be applied to new datasets (see methods) to help generate and validate new insights.

One important limitation is that we could only include patients who had undergone in-laboratory polysomnography, as trait estimates from home sleep apnea tests without electroencephalography have not yet been made available.^53^ Also, given the retrospective nature of the study we relied on the availability of adherence data in cloud databases. Thus, our results may not generalize to patients who did not undergo an in-laboratory polysomnography or whose adherence data were not retained. Furthermore, since this is an observational study, confounding and/or selection bias cannot be excluded.

Future studies should repeat the LPA procedure to assess reproducibility of the identified clusters, and apply the developed LPA model to new clinical datasets to validate the observed associations with adherence—and potentially other outcomes.^54^ The ongoing NICEPAP study (NCT05067088), a prospective real-world cohort examining the role of physiological traits in CPAP adherence and efficacy, could help replicate our current findings. Ultimately, however, interventional studies will be needed to determine the clinical value of “endotype” clusters for a precision medicine approach to OSA. For example, an ongoing trial testing the effect of the hypnotic eszopiclone on CPAP usage in patients who are struggling with adherence (TOP-CPAP, NCT05951023) may help clarify whether adherence in the “Low ArTH Driven” cluster can be improved with pharmacotherapy.

## Data Availability

All data produced in the present study are available upon reasonable request to the authors, approval by the UCSD IRB and a Data Use Agreement.

## CONCLUSION

In summary, while an existing trait-based linear model for predicting CPAP adherence did not perform well in our diverse clinical cohort, a latent profile analysis identified six distinct trait constellations, including a “Low ArTH Driven” cluster associated with persistently low adherence during the first three months of CPAP use. This non-linear approach appears promising to move from individual traits to actual endotypes, but external validation and interventional studies will be essential to determine its potential clinical value.

### Declaration of Generative AI and AI-assisted technologies in the writing process

During the preparation of this work the author CNS used OpenAI’s ChatGPT in order to edit the draft for clarity and conciseness. After using this tool/service, the authors reviewed and edited the content as needed and take full responsibility for the content of the publication.

## PRIOR ABSTRACT

Preliminary results of some of this work have been previously reported as an abstract and presented as a poster during the annual conference of the American Thoracic Society (ATS) in San Diego in May 2024. Schmickl CN, Harding C, Orr JE, et al. Going From Endotypical Traits to Actual Endotypes Using a Latent Profile Analysis. A110. NEW FRONTIERS IN SLEEP APNEA DIAGNOSIS AND THERAPIES: American Thoracic Society; 2024:A2979-A2979.

## AUTHOR CONTRIBUTIONS & GUARANTOR STATEMENT

All authors contributed substantially to the study conception and design (CNS, CH, AT, JEO, AZ, RLO, AM), data acquisition (CNS), analysis (CNS, CH, AT), and/or interpretation (all) of this study. CNS drafted the manuscript, and all authors revised it critically for intellectual content. All authors gave final approval of this version to be submitted.

CNS had full access to all of the data in the study and takes responsibility for the integrity of the data and the accuracy of the data analysis.

## ABBREVIATIONS

AHI: apnea hypopnea index
AIC: Akaike Information criterion
ArTH: arousal threshold
AUC: area under the curve
CAD: coronary artery disease
CI: confidence interval
CPAP: continuous positive airway pressure
IQR: interquartile range
IRB: institutional review board
(N)REM: (non-)rapid eye movement sleep
OSA: obstructive sleep apnea
SNOOzzzE: San Diego Multi-Outcome OSA Endophenotype cohort
UA: upper airway
VRA: ventilatory response to arousal

## ONLINE SUPPLEMENT

## E-Appendix 1. Development of the Trait Cluster (Endotype) model

Several traits contributing to the pathogenesis of obstructive sleep apnea (OSA) can be estimated from standard polysomnography. However, the thresholds at which these traits become pathophysiologically meaningful—and how they interact to drive disease expression—remain incompletely understood. Our objective was to use a latent profile analysis (LPA) to develop a data-driven clustering model of obstructive sleep apnea (OSA) endotypes based on key pathophysiological traits.

### METHODS

Gaussian finite mixture modeling (GMM, i.e., latent profile analysis), a model-based clustering method for continuous variables, was employed using a standard approach to identify distinct clusters (i.e., latent profiles) based on endotypical traits.^1,2^

#### Study Population

We used data from the SNOOzzzE cohort, which included 3,319 consecutive adults who underwent diagnostic in-laboratory polysomnography at the UCSD Sleep Center between 2017 and 2019. We excluded individuals without obstructive sleep apnea (OSA), those using supplemental oxygen during the study, and those for whom endotypical traits could not be estimated. Thus, the final cohort consisted of 2,367 patients which was randomly split into a training set (70%, *N*=1,655) and a test set (30%, *N*=712).

#### Measurements of Pathophysiological Traits (Indicators)

Mechanisms underlying OSA were estimated from raw signals obtained during NREM-sleep portions of the in-laboratory polysomnography using published techniques.^3–6^ To assess upper airway collapsibility we quantified Vpassive, which estimates the flow as a percentage of eupneic ventilation (%eupnea) at eupneic drive (lower Vpassive denotes worse collapsibility). To assess responsiveness of upper airway dilator muscles, we quantified Vactive (flow at elevated drive, specifically the level of the arousal threshold). Vcomp (Vactive-Vpassive) reflects the change in flow from passive to active conditions. Higher values of Vactive and Vcomp denote better upper airway dilator muscle function. Ventilatory instability (“loop gain”) was estimated as the magnitude of the ventilatory drive response to a prior reduction in ventilation in the time frame of 1 minute (loop gain 1, LG1) and the natural frequency (loop gain n, LGn; range 0 to infinity, higher levels of loop gain denote more instability). The arousal threshold (ArTH) was estimated as the level of respiratory drive causing arousals (lower values denoting easier arousability). Furthermore, the ventilatory response to arousal was estimated as the ventilatory overshoot (%eupnea) in response to arousals (greater overshoots may worsen sleep apnea)^7,8^, and circulatory delay was estimated as the latency between a drop in ventilation and a subsequent rise in chemical drive (principally the circulation time between the lung and chemoreceptors).^3^ Vpassive (Vpassive^T^ = 100-100*sqrt[1-Vpassive/100]) and Arousal Threshold (ArTH^T^ = 100+100*sqrt[ArTH/100-1]) were transformed for normality as in prior studies with transformed values used throughout.^9,10^

#### Data Setup

➢ Indicator selection: To reduce redundancy and capture distinct physiological mechanisms, we selected five traits as indicators for the latent profile analysis: upper airway collapsibility (Vpassive^T^), upper airway compensation (Vcomp), loop gain (LGn), arousal threshold (ArTH), and ventilatory response to arousal (VRA). Loop gain was represented by LGn rather than LG1 due to high correlation between the two (r = 0.76), and because LGn was less correlated with circulatory delay—a trait not included in the current model due to its less established role in OSA pathogenesis, but which may be added in future investigations. Similarly, Vcomp was selected over Vactive as it conceptually better reflects upper airway responsiveness. Of note, even though Vcomp is partially derived from Vpassive, the correlation between the two was minimal, supporting their joint inclusion in the model.
➢ Data processing: Included traits were standardized prior to analysis to facilitate model convergence and ensure comparability of trait variances.
➢ Sample Size Considerations: While our training set included 1,655 patients, a sample size >500 is generally considered adequate for model development.^2^

#### Model Selection & Validation

In the training set, we performed the LPA using the *mclust* package in R. GMMs were fit using 1 to 9 components across a range of possible variance-covariance structures (e.g. spherical, equal volume to ellipsoidal with varying volume, shape and orientation). Model selection was guided primarily by the Bayesian Information Criterion (BIC), with additional consideration of entropy (a measure of how well classes are separated; aiming for ∼0.8), minimum/maximum posterior probabilities, minimum profile sizes (at least 3%), bootstrapped likelihood ratio tests (BLRT; testing a k vs k-1 model, aiming for p<0.05). Moreover, particular emphasis was placed on ensuring that the resulting clusters were not only biologically interpretable but also qualitatively distinct from one another. Importantly, the optimal model was selected prior to linking the clusters to any outcome variables (e.g., adherence).^1,2^

Model validation in the test set proceeded in two steps:

1. Application of the training model to the test set: Cluster membership was assigned using the training model, and heatmaps were used to assess whether trait distributions across matched profiles are similar (to assess whether trait profiles and thus qualitative labels are stable).
2. Independent model development: A new model was developed in the test set using the same procedure as in the training set, and resulting profiles were compared for structural and biological similarity to the original clusters (to assess reproducibility/generalizability).

### RESULTS

The overall cohort was middle aged, had on average moderate OSA, and was highly diverse including 44% women, 45% reporting non-White race, and 24% reporting Hispanic/Latino ethnicity, with similar characteristics in the training and test set (**E-Table 1**).

Based on the BIC, the optimal LPA model had 6 clusters (technically “profiles”) with varying variances and covariances (VVV; **E-Figure 1**). Secondary metrics supported selection of the 6-cluster VVV model as well (**E-Table 2**): the average posterior classification probability reflected by the entropy of 0.78 was good (min = 0.73, max = 0.988), and the smallest cluster included 9.9% of the sample, while the BLRT was significant (P=0.01). Importantly, the 6-cluster VVV-solution also had qualitatively distinct, biologically meaningful profiles which were labeled based on the presumptive OSA-driving mechanisms (**E-Figure 2**).

Patients in the test set were then classified based on this training model, yielding similar profile sizes and trait distributions as in the training set (**E-Figure 2**). This consistency supports the stability of the identified clusters and the validity of their biological interpretations and applied labels.

When developing a new model in the test set, six clusters were again optimal. Five profiles were near-identical to the ones found in the training model, while the sixth profile partially replicated (**E-Figure 2**).

To allow future applications, a github repository was created that contains an end-to-end pipeline for applying the 6-cluster VVV LPA-model developed in the training set to new trait data (derived from PUPbeta): https://github.com/CSchmickl/SNOOzzzE_Endocluster_LPA_Pipeline

**E-Figure A1.**
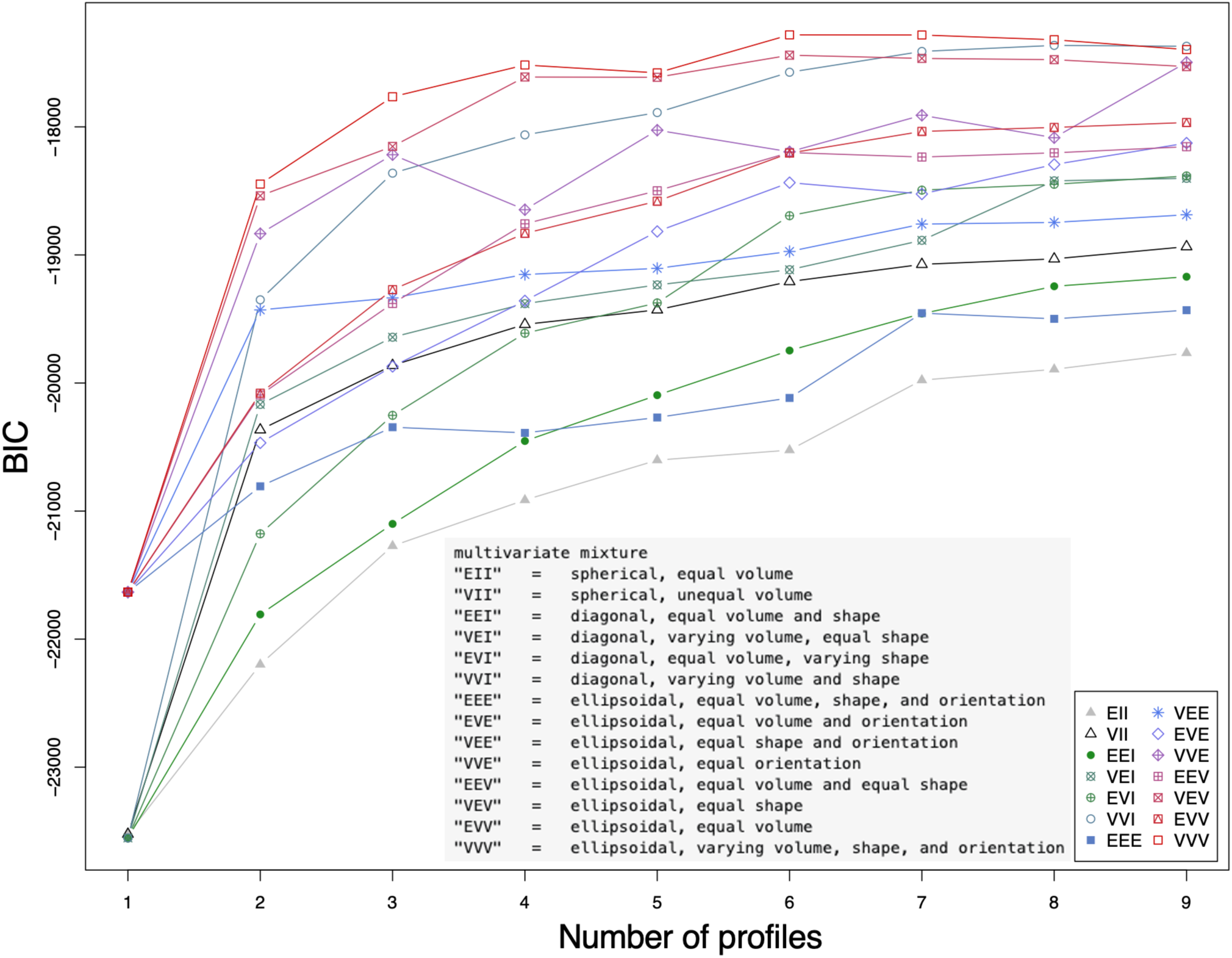
BICs for LPA-models with Different Variance/Covariances Structures and Number of Profiles.

**E-Figure A2.**
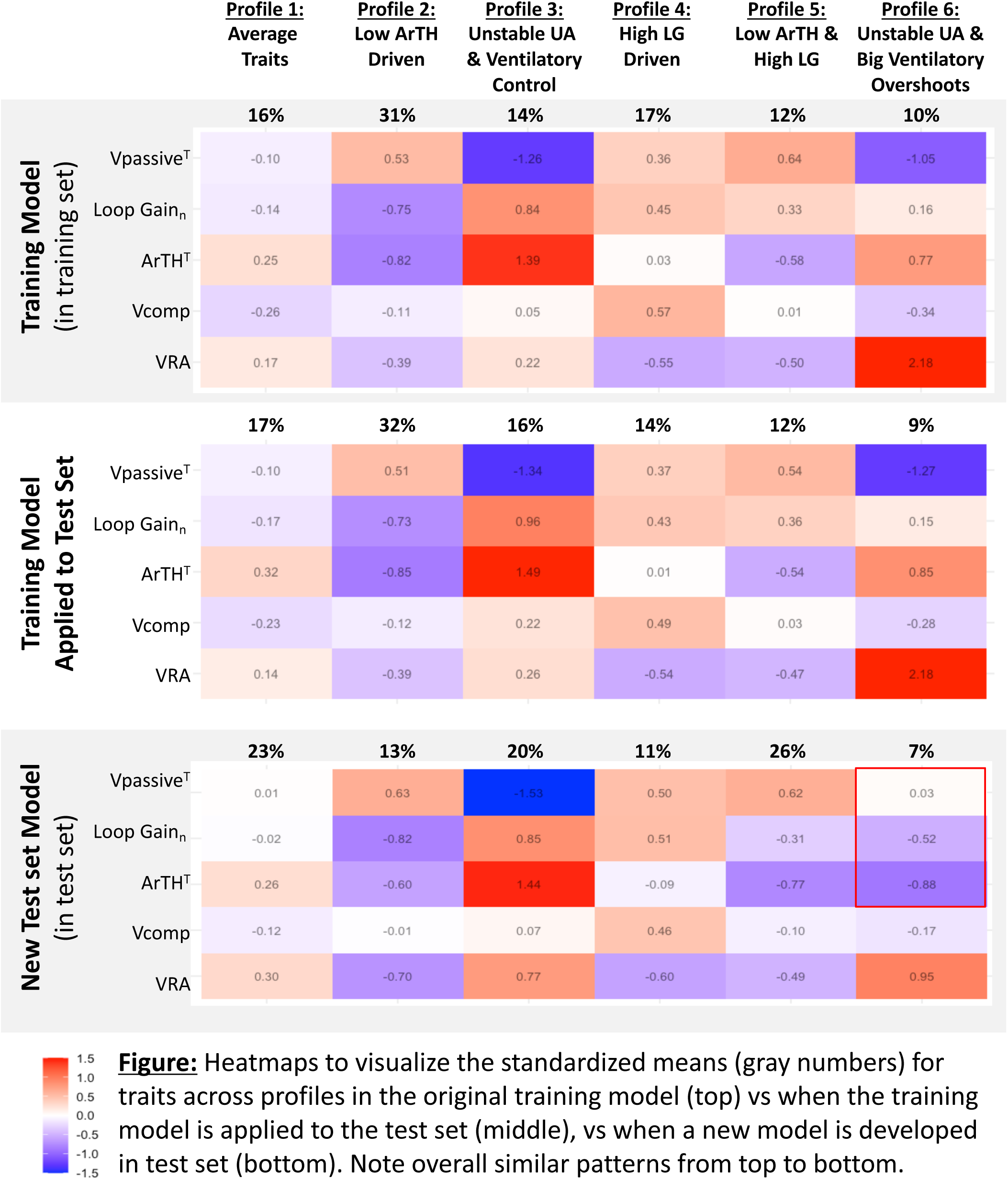
Characteristics of the Identified Trait Profiles (“Clusters”).

**E-Table A1.**
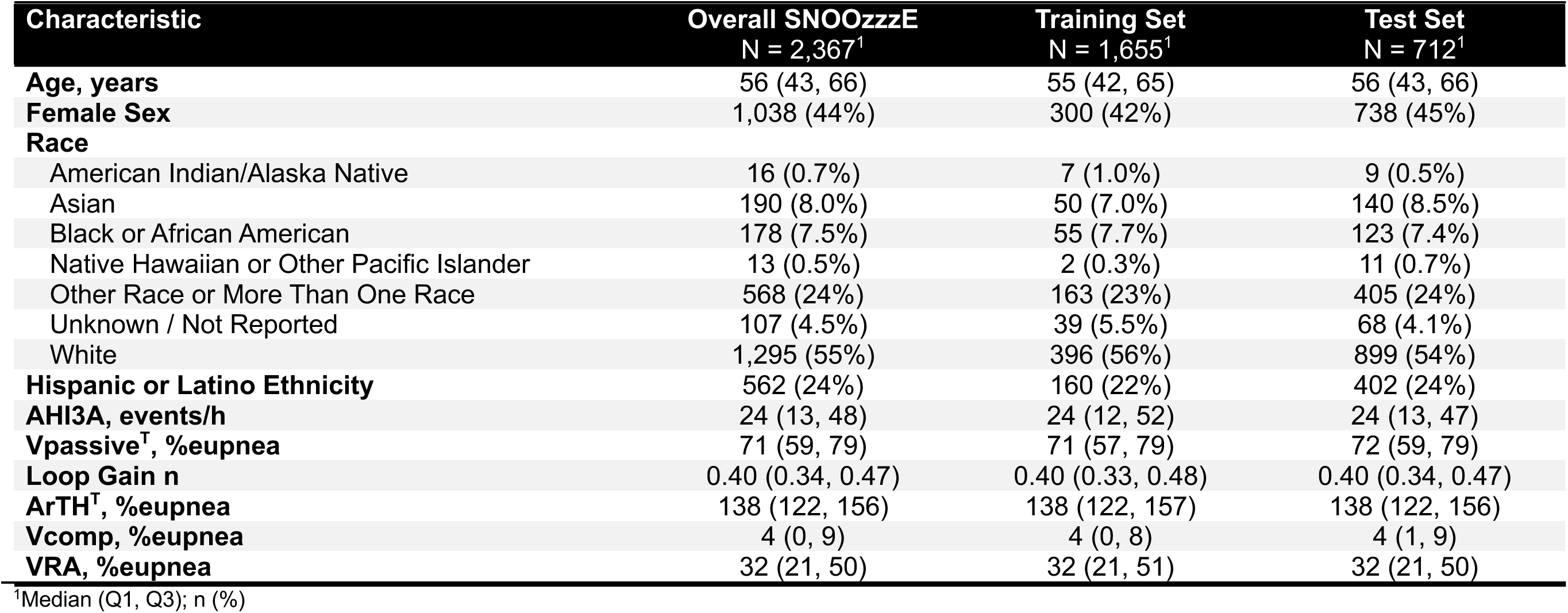
General Characteristics of the Training and Test Set Used for LPA.

**E-Table A2.** BIC and Secondary Metrics for VVV-Models with 1-9 Clusters (“Classes”).

| Classes | BIC | Entropy | prob_min | prob_max | n_min | n_max | BLRT_val | BLRT_p |
| --- | --- | --- | --- | --- | --- | --- | --- | --- |
| 1 | 21633.05 | 1.0000000 | 1.0000000 | 1.0000000 | 1.00000000 | 1.0000000 | NA | NA |
| 2 | 18446.66 | 0.8389134 | 0.9338185 | 0.9745121 | 0.39214502 | 0.6078550 | 3342.03256 | 0.00990099 |
| 3 | 17765.07 | 0.8057988 | 0.8623604 | 0.9376287 | 0.28036254 | 0.4066465 | 837.23146 | 0.00990099 |
| 4 | 17517.03 | 0.7745028 | 0.8008786 | 0.9546164 | 0.19335347 | 0.4006042 | 403.68733 | 0.00990099 |
| 5 | 17578.45 | 0.7540081 | 0.7469883 | 0.9036175 | 0.12990937 | 0.3039275 | 94.22079 | 0.00990099 |
| 6 | 17281.48 | 0.7842486 | 0.7335161 | 0.9884402 | 0.09909366 | 0.3087613 | 452.61411 | 0.00990099 |
| 7 | 17282.69 | 0.7893826 | 0.7269581 | 0.9810426 | 0.08700906 | 0.3057402 | 154.43242 | 0.00990099 |
| 8 | 17319.79 | 0.7749431 | 0.6727668 | 0.9872748 | 0.06948640 | 0.2549849 | 118.53961 | 0.00990099 |
| 9 | 17396.50 | 0.7762095 | 0.6636874 | 0.9856499 | 0.03685801 | 0.2513595 | 78.93351 | 0.00990099 |

**E-Figure 1.**
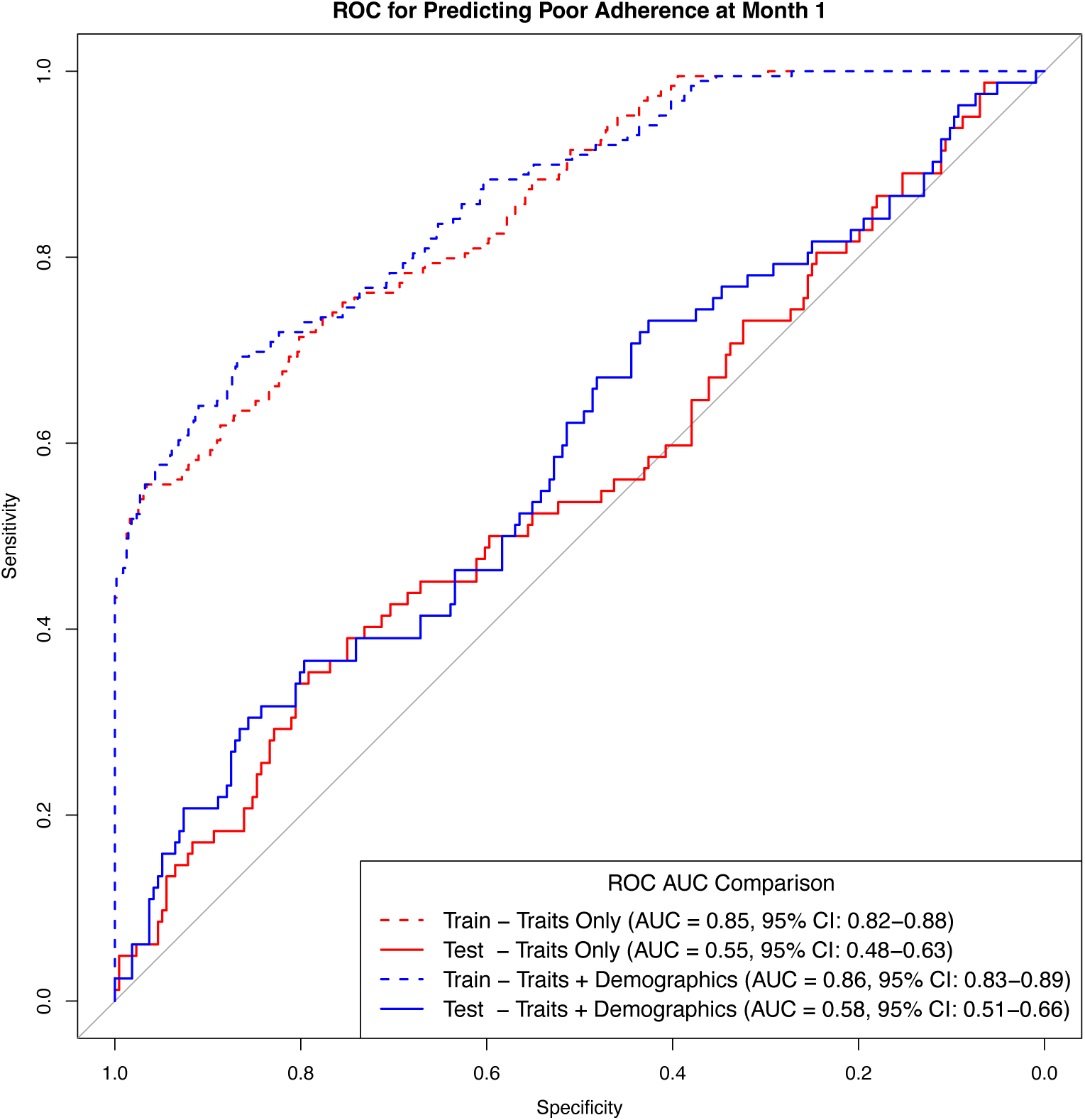
ROCs of the De Novo Linear Trait Models. Trait only (red) and Trait + Demographics (blue) model performed both similarly well in the training set (dashed lines), but performed both poorly in the test set (solid lines). For model details see E-Table 3.

**E-Figure 2.**
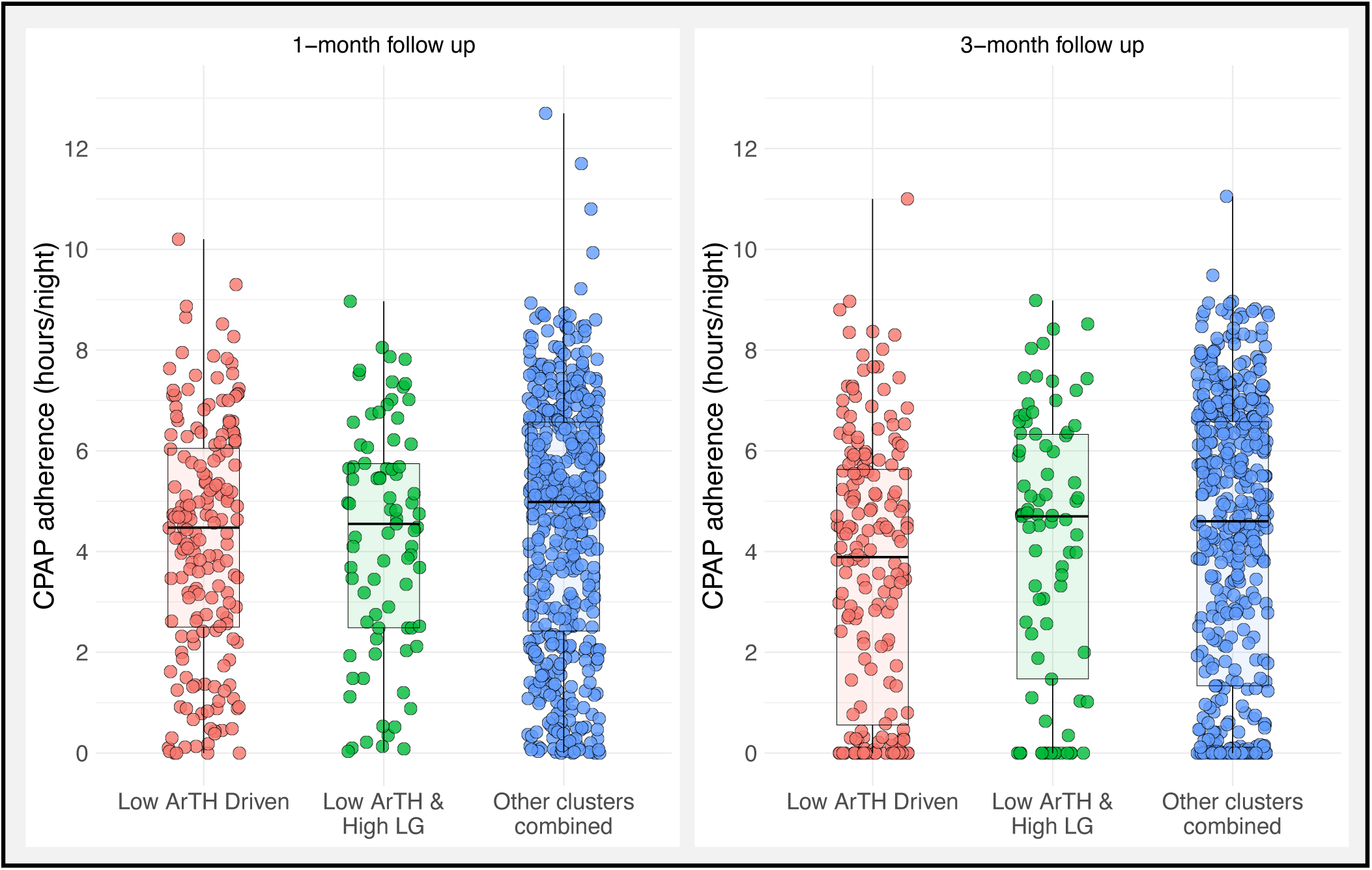
Boxplots of CPAP Adherence at 1 & 3 Months in the low-ArTH vs Other Clusters.

**E-Table 1.** Details of the *De Novo* Linear Trait Model. Trait variables were mean-centered using the sample means, thus the intercept reflects the predicted CPAP adherence for someone with mean traits.

|  | Traits Only |  |  | Traits + Age, Sex, Race, Ethnicity |  |  |
| --- | --- | --- | --- | --- | --- | --- |
|  | Beta | 95% CI<sup>1</sup> | p-value | Beta | 95% CI<sup>1</sup> | p-value |
| Intercept | 4.9 | 4.6, 5.2 | <0.001 | 3.6 | 2.5, 4.7 | <0.001 |
| Month | -0.22 | -0.31, -0.13 | <0.001 | -0.22 | -0.31, -0.13 | <0.001 |
| ArTH<sup>T</sup>, %eupnea | 0.002 | -0.01, 0.01 | 0.7 | -0.005 | -0.01, 0.01 | 0.4 |
| LG1 | 1.6 | 0.25, 3.0 | 0.021 | 1.3 | -0.03, 2.7 | 0.055 |
| Vcomp, %eupnea | -0.01 | -0.02, 0.00 | 0.059 | -0.01 | -0.02, 0.00 | 0.044 |
| Vpassive<sup>T</sup>, %eupnea | -0.01 | -0.02, 0.01 | 0.3 | -0.01 | -0.02, 0.01 | 0.3 |
| LG1 * Vpassive<sup>T</sup> | 0.03 | -0.01, 0.07 | 0.13 | 0.02 | -0.03, 0.06 | 0.5 |

<sup>1</sup>CI = Confidence Interval

**E-Table 2.**
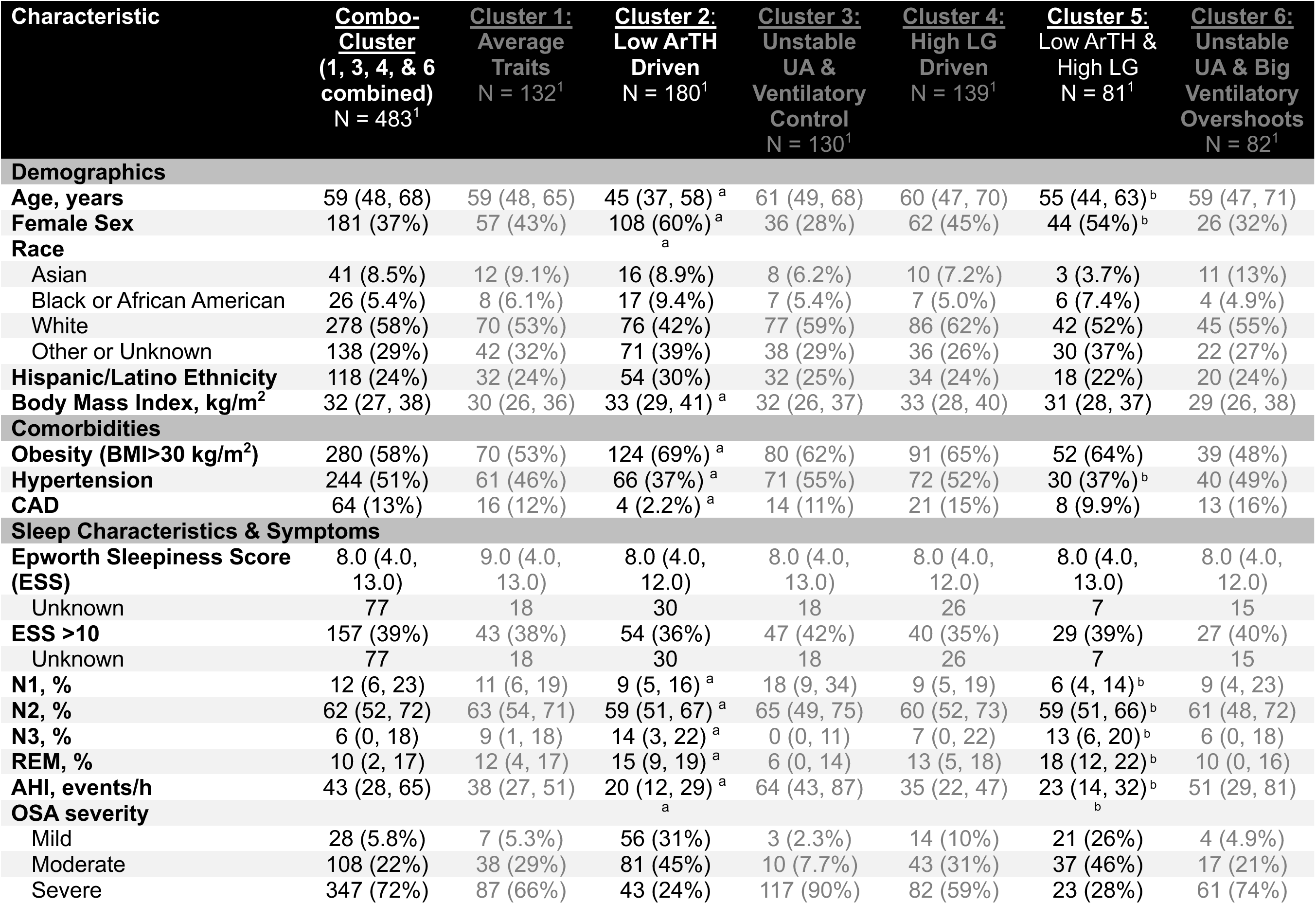
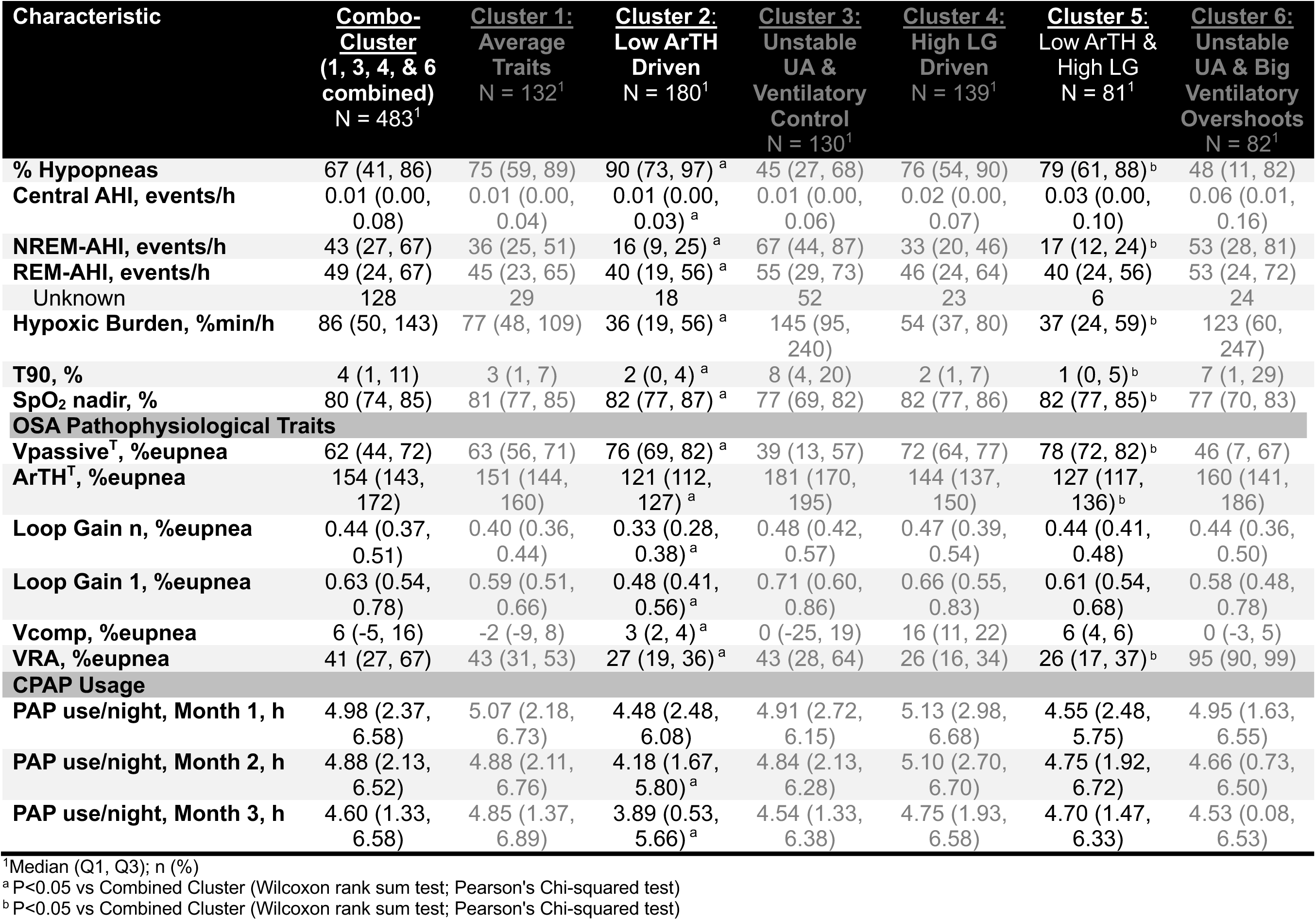
General Characteristics of Endotype Clusters. For this analysis, clusters characterized by a “low ArTH” (cluster 1 and 2) were each contrasted against the combination of all clusters (reference).

| **Characteristic** | **Combo-Cluster**<br>**(1, 3, 4, & 6**<br>**combined)**<br>**N = 483<sup>1</sup>** | **Cluster 1:**<br>**Average**<br>**Traits**<br>**N = 132<sup>1</sup>** | **Cluster 2:**<br>**Low ArTH**<br>**Driven**<br>**N = 180<sup>1</sup>** | **Cluster 3:**<br>**Unstable**<br>**UA &**<br>**Ventilatory**<br>**Control**<br>**N = 130<sup>1</sup>** | **Cluster 4:**<br>**High LG**<br>**Driven**<br>**N = 139<sup>1</sup>** | **Cluster 5:**<br>**Low ArTH &**<br>**High LG**<br>**N = 81<sup>1</sup>** | **Cluster 6:**<br>**Unstable**<br>**UA & Big**<br>**Ventilatory**<br>**Overshoots**<br>**N = 82<sup>1</sup>** |
| --- | --- | --- | --- | --- | --- | --- | --- |
| **Demographics** |  |  |  |  |  |  |  |
| **Age, years** | 59 (48, 68) | 59 (48, 65) | 45 (37, 58) <sup>a</sup> | 61 (49, 68) | 60 (47, 70) | 55 (44, 63) <sup>b</sup> | 59 (47, 71) |
| **Female Sex** | 181 (37%) | 57 (43%) | 108 (60%) <sup>a</sup> | 36 (28%) | 62 (45%) | 44 (54%) <sup>b</sup> | 26 (32%) |
| **Race** |  |  |  |  |  |  |  |
| Asian | 41 (8.5%) | 12 (9.1%) | 16 (8.9%) | 8 (6.2%) | 10 (7.2%) | 3 (3.7%) | 11 (13%) |
| Black or African American | 26 (5.4%) | 8 (6.1%) | 17 (9.4%) | 7 (5.4%) | 7 (5.0%) | 6 (7.4%) | 4 (4.9%) |
| White | 278 (58%) | 70 (53%) | 76 (42%) | 77 (59%) | 86 (62%) | 42 (52%) | 45 (55%) |
| Other or Unknown | 138 (29%) | 42 (32%) | 71 (39%) | 38 (29%) | 36 (26%) | 30 (37%) | 22 (27%) |
| **Hispanic/Latino Ethnicity** | 118 (24%) | 32 (24%) | 54 (30%) | 32 (25%) | 34 (24%) | 18 (22%) | 20 (24%) |
| **Body Mass Index, kg/m<sup>2</sup>** | 32 (27, 38) | 30 (26, 36) | 33 (29, 41) <sup>a</sup> | 32 (26, 37) | 33 (28, 40) | 31 (28, 37) | 29 (26, 38) |
| **Comorbidities** |  |  |  |  |  |  |  |
| **Obesity (BMI>30 kg/m<sup>2</sup>)** | 280 (58%) | 70 (53%) | 124 (69%) <sup>a</sup> | 80 (62%) | 91 (65%) | 52 (64%) | 39 (48%) |
| **Hypertension** | 244 (51%) | 61 (46%) | 66 (37%) <sup>a</sup> | 71 (55%) | 72 (52%) | 30 (37%) <sup>b</sup> | 40 (49%) |
| **CAD** | 64 (13%) | 16 (12%) | 4 (2.2%) <sup>a</sup> | 14 (11%) | 21 (15%) | 8 (9.9%) | 13 (16%) |
| **Sleep Characteristics & Symptoms** |  |  |  |  |  |  |  |
| **Epworth Sleepiness Score (ESS)** | 8.0 (4.0, 13.0) | 9.0 (4.0, 13.0) | 8.0 (4.0, 12.0) | 8.0 (4.0, 13.0) | 8.0 (4.0, 12.0) | 8.0 (4.0, 13.0) | 8.0 (4.0, 12.0) |
| Unknown | 77 | 18 | 30 | 18 | 26 | 7 | 15 |
| **ESS >10** | 157 (39%) | 43 (38%) | 54 (36%) | 47 (42%) | 40 (35%) | 29 (39%) | 27 (40%) |
| Unknown | 77 | 18 | 30 | 18 | 26 | 7 | 15 |
| **N1, %** | 12 (6, 23) | 11 (6, 19) | 9 (5, 16) <sup>a</sup> | 18 (9, 34) | 9 (5, 19) | 6 (4, 14) <sup>b</sup> | 9 (4, 23) |
| **N2, %** | 62 (52, 72) | 63 (54, 71) | 59 (51, 67) <sup>a</sup> | 65 (49, 75) | 60 (52, 73) | 59 (51, 66) <sup>b</sup> | 61 (48, 72) |
| **N3, %** | 6 (0, 18) | 9 (1, 18) | 14 (3, 22) <sup>a</sup> | 0 (0, 11) | 7 (0, 22) | 13 (6, 20) <sup>b</sup> | 6 (0, 18) |
| **REM, %** | 10 (2, 17) | 12 (4, 17) | 15 (9, 19) <sup>a</sup> | 6 (0, 14) | 13 (5, 18) | 18 (12, 22) <sup>b</sup> | 10 (0, 16) |
| **AHI, events/h** | 43 (28, 65) | 38 (27, 51) | 20 (12, 29) <sup>a</sup> | 64 (43, 87) | 35 (22, 47) | 23 (14, 32) <sup>b</sup> | 51 (29, 81) |
| **OSA severity** |  |  |  |  |  |  |  |
| Mild | 28 (5.8%) | 7 (5.3%) | 56 (31%) | 3 (2.3%) | 14 (10%) | 21 (26%) | 4 (4.9%) |
| Moderate | 108 (22%) | 38 (29%) | 81 (45%) | 10 (7.7%) | 43 (31%) | 37 (46%) | 17 (21%) |
| Severe | 347 (72%) | 87 (66%) | 43 (24%) | 117 (90%) | 82 (59%) | 23 (28%) | 61 (74%) |

| Characteristic | **Combo-Cluster**<br>(1, 3, 4, & 6 combined)<br>N = 483<sup>1</sup> | **Cluster 1:**<br>Average Traits<br>N = 132<sup>1</sup> | **Cluster 2:**<br>Low ArTH Driven<br>N = 180<sup>1</sup> | **Cluster 3:**<br>Unstable UA & Ventilatory Control<br>N = 130<sup>1</sup> | **Cluster 4:**<br>High LG Driven<br>N = 139<sup>1</sup> | **Cluster 5:**<br>Low ArTH & High LG<br>N = 81<sup>1</sup> | **Cluster 6:**<br>Unstable UA & Big Ventilatory Overshoots<br>N = 82<sup>1</sup> |
| --- | --- | --- | --- | --- | --- | --- | --- |
| % Hypopneas | 67 (41, 86) | 75 (59, 89) | 90 (73, 97)<sup>a</sup> | 45 (27, 68) | 76 (54, 90) | 79 (61, 88)<sup>b</sup> | 48 (11, 82) |
| Central AHI, events/h | 0.01 (0.00, 0.08) | 0.01 (0.00, 0.04) | 0.01 (0.00, 0.03)<sup>a</sup> | 0.01 (0.00, 0.06) | 0.02 (0.00, 0.07) | 0.03 (0.00, 0.10) | 0.06 (0.01, 0.16) |
| NREM-AHI, events/h | 43 (27, 67) | 36 (25, 51) | 16 (9, 25)<sup>a</sup> | 67 (44, 87) | 33 (20, 46) | 17 (12, 24)<sup>b</sup> | 53 (28, 81) |
| REM-AHI, events/h | 49 (24, 67) | 45 (23, 65) | 40 (19, 56)<sup>a</sup> | 55 (29, 73) | 46 (24, 64) | 40 (24, 56) | 53 (24, 72) |
| Unknown | 128 | 29 | 18 | 52 | 23 | 6 | 24 |
| Hypoxic Burden, %min/h | 86 (50, 143) | 77 (48, 109) | 36 (19, 56)<sup>a</sup> | 145 (95, 240) | 54 (37, 80) | 37 (24, 59)<sup>b</sup> | 123 (60, 247) |
| T90, % | 4 (1, 11) | 3 (1, 7) | 2 (0, 4)<sup>a</sup> | 8 (4, 20) | 2 (1, 7) | 1 (0, 5)<sup>b</sup> | 7 (1, 29) |
| SpO<sub>2</sub> nadir, % | 80 (74, 85) | 81 (77, 85) | 82 (77, 87)<sup>a</sup> | 77 (69, 82) | 82 (77, 86) | 82 (77, 85)<sup>b</sup> | 77 (70, 83) |
| **OSA Pathophysiological Traits** |  |  |  |  |  |  |  |
| Vpassive<sup>T</sup>, %eupnea | 62 (44, 72) | 63 (56, 71) | 76 (69, 82)<sup>a</sup> | 39 (13, 57) | 72 (64, 77) | 78 (72, 82)<sup>b</sup> | 46 (7, 67) |
| ArTH<sup>T</sup>, %eupnea | 154 (143, 172) | 151 (144, 160) | 121 (112, 127)<sup>a</sup> | 181 (170, 195) | 144 (137, 150) | 127 (117, 136)<sup>b</sup> | 160 (141, 186) |
| Loop Gain n, %eupnea | 0.44 (0.37, 0.51) | 0.40 (0.36, 0.44) | 0.33 (0.28, 0.38)<sup>a</sup> | 0.48 (0.42, 0.57) | 0.47 (0.39, 0.54) | 0.44 (0.41, 0.48) | 0.44 (0.36, 0.50) |
| Loop Gain 1, %eupnea | 0.63 (0.54, 0.78) | 0.59 (0.51, 0.66) | 0.48 (0.41, 0.56)<sup>a</sup> | 0.71 (0.60, 0.86) | 0.66 (0.55, 0.83) | 0.61 (0.54, 0.68) | 0.58 (0.48, 0.78) |
| Vcomp, %eupnea | 6 (-5, 16) | -2 (-9, 8) | 3 (2, 4)<sup>a</sup> | 0 (-25, 19) | 16 (11, 22) | 6 (4, 6) | 0 (-3, 5) |
| VRA, %eupnea | 41 (27, 67) | 43 (31, 53) | 27 (19, 36)<sup>a</sup> | 43 (28, 64) | 26 (16, 34) | 26 (17, 37)<sup>b</sup> | 95 (90, 99) |
| **CPAP Usage** |  |  |  |  |  |  |  |
| PAP use/night, Month 1, h | 4.98 (2.37, 6.58) | 5.07 (2.18, 6.73) | 4.48 (2.48, 6.08) | 4.91 (2.72, 6.15) | 5.13 (2.98, 6.68) | 4.55 (2.48, 5.75) | 4.95 (1.63, 6.55) |
| PAP use/night, Month 2, h | 4.88 (2.13, 6.52) | 4.88 (2.11, 6.76) | 4.18 (1.67, 5.80)<sup>a</sup> | 4.84 (2.13, 6.28) | 5.10 (2.70, 6.70) | 4.75 (1.92, 6.72) | 4.66 (0.73, 6.50) |
| PAP use/night, Month 3, h | 4.60 (1.33, 6.58) | 4.85 (1.37, 6.89) | 3.89 (0.53, 5.66)<sup>a</sup> | 4.54 (1.33, 6.38) | 4.75 (1.93, 6.58) | 4.70 (1.47, 6.33) | 4.53 (0.08, 6.53) |

<sup>1</sup>Median (Q1, Q3); n (%)
<sup>a</sup>P<0.05 vs Combined Cluster (Wilcoxon rank sum test; Pearson's Chi-squared test)
<sup>b</sup>P<0.05 vs Combined Cluster (Wilcoxon rank sum test; Pearson's Chi-squared test)

## REFERENCES

1. Jordan AS, White DP, Owens RL, et al. The effect of increased genioglossus activity and end-expiratory lung volume on pharyngeal collapse. Journal of applied physiology (Bethesda, Md.: 1985). 2010;109(2):469–475.

2. Benjafield AV, Ayas NT, Eastwood PR, et al. Estimation of the global prevalence and burden of obstructive sleep apnoea: a literature-based analysis. The Lancet. Respiratory medicine. 2019;7(8):687–698.

3. Peppard PE, Young T, Barnet JH, Palta M, Hagen EW, Hla KM. Increased prevalence of sleep-disordered breathing in adults. American journal of epidemiology. 2013;177(9):1006–1014.

4. Chervin RD. Sleepiness, fatigue, tiredness, and lack of energy in obstructive sleep apnea. Chest. 2000;118(2):372–379.

5. Jenkinson C, Davies RJ, Mullins R, Stradling JR. Comparison of therapeutic and subtherapeutic nasal continuous positive airway pressure for obstructive sleep apnoea: a randomised prospective parallel trial. *Lancet (London*, England). 1999;353(9170):2100–2105.

6. Teran-Santos J, Jimenez-Gomez A, Cordero-Guevara J. The association between sleep apnea and the risk of traffic accidents. Cooperative Group Burgos-Santander. The New England journal of medicine. 1999;340(11):847–851.

7. Tregear S, Reston J, Schoelles K, Phillips B. Obstructive sleep apnea and risk of motor vehicle crash: systematic review and meta-analysis. Journal of clinical sleep medicine: JCSM: official publication of the American Academy of Sleep Medicine. 2009;5(6):573–581.

8. Lavie P, Herer P, Hoffstein V. Obstructive sleep apnoea syndrome as a risk factor for hypertension: population study. BMJ (Clinical research ed.). 2000;320(7233):479–482.

9. Marin JM, Agusti A, Villar I, et al. Association between treated and untreated obstructive sleep apnea and risk of hypertension. Jama. 2012;307(20):2169–2176.

10. Gottlieb DJ, Yenokyan G, Newman AB, et al. Prospective study of obstructive sleep apnea and incident coronary heart disease and heart failure: the sleep heart health study. Circulation. 2010;122(4):352–360.

11. Young T, Finn L, Peppard PE, et al. Sleep disordered breathing and mortality: eighteen-year follow-up of the Wisconsin sleep cohort. Sleep. 2008;31(8):1071–1078.

12. Sullivan CE, Issa FG, Berthon-Jones M, Eves L. Reversal of obstructive sleep apnoea by continuous positive airway pressure applied through the nares. *Lancet (London*, England). 1981;1(8225):862–865.

13. Weaver TE, Grunstein RR. Adherence to continuous positive airway pressure therapy: the challenge to effective treatment. Proceedings of the American Thoracic Society. 2008;5(2):173–178.

14. Rotenberg BW, Murariu D, Pang KP. Trends in CPAP adherence over twenty years of data collection: a flattened curve. J Otolaryngol Head Neck Surg. 2016;45(1):43.

15. Cistulli PA, Armitstead J, Pepin JL, et al. Short-term CPAP adherence in obstructive sleep apnea: a big data analysis using real world data.59:114–116.

16. Drager LF, Malhotra A, Yan Y, et al. Adherence with positive airway pressure therapy for obstructive sleep apnea in developing vs. developed countries: a big data study. Journal of clinical sleep medicine: JCSM: official publication of the American Academy of Sleep Medicine. 2021;17(4):703–709.

17. Crawford MR, Espie CA, Bartlett DJ, Grunstein RR. Integrating psychology and medicine in CPAP adherence--new concepts? Sleep medicine reviews. 2014;18(2):123–139.

18. Sawyer AM, Gooneratne NS, Marcus CL, Ofer D, Richards KC, Weaver TE. A systematic review of CPAP adherence across age groups: clinical and empiric insights for developing CPAP adherence interventions. Sleep medicine reviews. 2011;15(6):343–356.

19. Eckert DJ, White DP, Jordan AS, Malhotra A, Wellman A. Defining phenotypic causes of obstructive sleep apnea. Identification of novel therapeutic targets. American journal of respiratory and critical care medicine. 2013;188(8):996–1004.

20. Wellman A, Edwards BA, Sands SA, et al. A simplified method for determining phenotypic traits in patients with obstructive sleep apnea. Journal of applied physiology (Bethesda, Md.: 1985). 2013;114(7):911–922.

21. Tolbert TM, Schmickl CN, Gell LK, et al. Research Priorities for Translating Endophenotyping of Adult Obstructive Sleep Apnea to the Clinic. An Official American Thoracic Society Research Statement. American journal of respiratory and critical care medicine. 2025.

22. Edwards BA, Eckert DJ, McSharry DG, et al. Clinical predictors of the respiratory arousal threshold in patients with obstructive sleep apnea. American journal of respiratory and critical care medicine. 2014;190(11):1293–1300.

23. Zinchuk A, Edwards BA, Jeon S, et al. Prevalence, Associated Clinical Features, and Impact on Continuous Positive Airway Pressure Use of a Low Respiratory Arousal Threshold Among Male United States Veterans With Obstructive Sleep Apnea. Journal of clinical sleep medicine: JCSM: official publication of the American Academy of Sleep Medicine. 2018;14(5):809–817.

24. Zinchuk AV, Chu JH, Liang J, et al. Physiological Traits and Adherence to Therapy of Sleep Apnea in Individuals with Coronary Artery Disease. American journal of respiratory and critical care medicine. 2021.

25. Harding CD, Fuentes AL, Malhotra A. Tackling obstructive sleep apnea with pharmacotherapeutics: expert guidance. Expert opinion on pharmacotherapy. 2024;25(8):1019–1026.

26. Younes M, Schwab R. Con: can physiological risk factors for obstructive sleep apnea be determined by analysis of data obtained from routine polysomnography? Sleep. 2023;46(5).

27. Sands SA, Edwards BA. Pro: can physiological risk factors for obstructive sleep apnea be determined by analysis of data obtained from routine polysomnography? Sleep. 2023;46(5).

28. Terrill PI, Edwards BA, Nemati S, et al. Quantifying the ventilatory control contribution to sleep apnoea using polysomnography. The European respiratory journal. 2015;45(2):408–418.

29. Sands SA, Edwards BA, Terrill PI, et al. Phenotyping Pharyngeal Pathophysiology Using Polysomnography in Patients with Obstructive Sleep Apnea. American journal of respiratory and critical care medicine. 2018;1(9):1187–1197.

30. Vena D, Taranto-Montemurro L, Azarbarzin A, et al. Clinical polysomnographic methods for estimating pharyngeal collapsibility in obstructive sleep apnea. Sleep. 2022;45(6).

31. Sands SA, Terrill PI, Edwards BA, et al. Quantifying the Arousal Threshold Using Polysomnography in Obstructive Sleep Apnea. Sleep. 2018;41(1).

32. Edwards BA, Connolly JG, Campana LM, et al. Acetazolamide attenuates the ventilatory response to arousal in patients with obstructive sleep apnea. Sleep. 2013;36(2):281–285.

33. Jordan AS, McEvoy RD, Edwards JK, et al. The influence of gender and upper airway resistance on the ventilatory response to arousal in obstructive sleep apnoea in humans. The Journal of physiology. 2004;558(Pt 3):993–1004.

34. Sands SA, Edwards BA, Terrill PI, et al. Identifying obstructive sleep apnoea patients responsive to supplemental oxygen therapy. The European respiratory journal. 2018;52(3).

35. O’Driscoll DM, Landry SA, Pham J, et al. The physiological phenotype of obstructive sleep apnea differs between Caucasian and Chinese patients. Sleep. 2019;42(11).

36. Sinha P, Calfee CS, Delucchi KL. Practitioner’s Guide to Latent Class Analysis: Methodological Considerations and Common Pitfalls. Critical care medicine. 2021;49(1):e63–e79.

37. Spurk D, Hirschi A, Wang M, Valero D, Kauffeld S. Latent profile analysis: A review and “how to” guide of its application within vocational behavior research. Journal of Vocational Behavior. 2020;120:103445.

38. Schmickl CN, Harding C, Orr JE, et al. Going From Endotypical Traits to Actual Endotypes Using a Latent Profile Analysis. A110. NEW FRONTIERS IN SLEEP APNEA DIAGNOSIS AND THERAPIES: American Thoracic Society; 2024:A2979–A2979.

39. Hsu N, Zeidler MR, Ryden AM, Fung CH. Racial disparities in positive airway pressure therapy adherence among veterans with obstructive sleep apnea. Journal of clinical sleep medicine: JCSM: official publication of the American Academy of Sleep Medicine. 2020;16(8):1249–1254.

40. Borker PV, Carmona E, Essien UR, et al. Neighborhoods with Greater Prevalence of Minority Residents Have Lower Continuous Positive Airway Pressure Adherence. American journal of respiratory and critical care medicine. 2021;204(3):339–346.

41. Thomson LDJ, Landry SA, Singleton R, et al. The Effect of Hypopnea Scoring on the Arousal Threshold in Patients with Obstructive Sleep Apnea. American journal of respiratory and critical care medicine. 2020.

42. Haba-Rubio J, Sforza E, Weiss T, Schroder C, Krieger J. Effect of CPAP treatment on inspiratory arousal threshold during NREM sleep in OSAS. Sleep & breathing = Schlaf & Atmung. 2005;9(1):12–19.

43. Loewen A, Ostrowski M, Laprairie J, et al. Determinants of ventilatory instability in obstructive sleep apnea: inherent or acquired? Sleep. 2009;32(10):1355–1365.

44. Orr JE, Edwards BA, Malhotra A. CrossTalk opposing view: Loop gain is not a consequence of obstructive sleep apnoea. The Journal of physiology. 2014;592(14):2903–2905.

45. Patil SP, Ayappa IA, Caples SM, Kimoff RJ, Patel SR, Harrod CG. Treatment of Adult Obstructive Sleep Apnea With Positive Airway Pressure: An American Academy of Sleep Medicine Systematic Review, Meta-Analysis, and GRADE Assessment. Journal of clinical sleep medicine: JCSM: official publication of the American Academy of Sleep Medicine. 2019;15(2):301–334.

46. Patel SR, Bakker JP, Stitt CJ, Aloia MS, Nouraie SM. Age and Sex Disparities in Adherence to CPAP. Chest. 2021;159(1):382–389.

47. Anderson GP. Endotyping asthma: new insights into key pathogenic mechanisms in a complex, heterogeneous disease. *Lancet (London*, England). 2008;372(9643):1107–1119.

48. Mazzotti DR, Keenan BT, Lim DC, Gottlieb DJ, Kim J, Pack AI. Symptom Subtypes of Obstructive Sleep Apnea Predict Incidence of Cardiovascular Outcomes. American journal of respiratory and critical care medicine. 2019;200(4):493–506.

49. Pien GW, Ye L, Keenan BT, et al. Changing Faces of Obstructive Sleep Apnea: Treatment Effects by Cluster Designation in the Icelandic Sleep Apnea Cohort. Sleep. 2018;41(3).

50. Ye L, Pien GW, Ratcliffe SJ, et al. The different clinical faces of obstructive sleep apnoea: a cluster analysis. The European respiratory journal. 2014;44(6):1600–1607.

51. Cheng WJ, Finnsson E, Arnardóttir E, Ágústsson JS, Sands SA, Hang LW. Relationship between Symptom Profiles and Endotypes among Patients with Obstructive Sleep Apnea: A Latent Class Analysis. Annals of the American Thoracic Society. 2023;20(9):1337–1344.

52. Landry SA, Joosten SA, Thomson LDJ, et al. Effect of Hypopnea Scoring Criteria on Noninvasive Assessment of Loop Gain and Surgical Outcome Prediction. Annals of the American Thoracic Society. 2019;17(4):484–491.

53. Orr JE, Sands SA, Edwards BA, et al. Measuring Loop Gain via Home Sleep Testing in Patients with Obstructive Sleep Apnea. American journal of respiratory and critical care medicine. 2018;197(10):1353–1355.

54. Edwards BA, Redline S, Sands SA, Owens RL. More Than the Sum of the Respiratory Events: Personalized Medicine Approaches for Obstructive Sleep Apnea. American journal of respiratory and critical care medicine. 2019;200(6):691–703.

## Supplemental References

1. Spurk D, Hirschi A, Wang M, Valero D, Kauffeld S. Latent profile analysis: A review and “how to” guide of its application within vocational behavior research. Journal of Vocational Behavior. 2020;120:103445.

2. Sinha P, Calfee CS, Delucchi KL. Practitioner’s Guide to Latent Class Analysis: Methodological Considerations and Common Pitfalls. Critical care medicine. 2021;49(1):e63–e79.

3. Terrill PI, Edwards BA, Nemati S, et al. Quantifying the ventilatory control contribution to sleep apnoea using polysomnography. The European respiratory journal. 2015;45(2):408–418.

4. Sands SA, Edwards BA, Terrill PI, et al. Phenotyping Pharyngeal Pathophysiology Using Polysomnography in Patients with Obstructive Sleep Apnea. American journal of respiratory and critical care medicine. 2018;1(9):1187–1197.

5. Vena D, Taranto-Montemurro L, Azarbarzin A, et al. Clinical polysomnographic methods for estimating pharyngeal collapsibility in obstructive sleep apnea. Sleep. 2022;45(6).

6. Sands SA, Terrill PI, Edwards BA, et al. Quantifying the Arousal Threshold Using Polysomnography in Obstructive Sleep Apnea. Sleep. 2018;41(1).

7. Edwards BA, Connolly JG, Campana LM, et al. Acetazolamide attenuates the ventilatory response to arousal in patients with obstructive sleep apnea. Sleep. 2013;36(2):281–285.

8. Jordan AS, McEvoy RD, Edwards JK, et al. The influence of gender and upper airway resistance on the ventilatory response to arousal in obstructive sleep apnoea in humans. The Journal of physiology. 2004;558(Pt 3):993–1004.

9. Sands SA, Edwards BA, Terrill PI, et al. Identifying obstructive sleep apnoea patients responsive to supplemental oxygen therapy. The European respiratory journal. 2018;52(3).

10. O’Driscoll DM, Landry SA, Pham J, et al. The physiological phenotype of obstructive sleep apnea differs between Caucasian and Chinese patients. Sleep. 2019;42(11).

